# A Novel Playbook for Pragmatic Trial Operations to Monitor and Evaluate Ambient Artificial Intelligence in Clinical Practice

**DOI:** 10.1101/2024.12.27.24319685

**Authors:** Majid Afshar, Felice Resnik, Mary Ryan Baumann, Josie Hintzke, Kayla Lemmon, Anne Gravel Sullivan, Tina Shah, Anthony Stordalen, Michael Oberst, Jason Dambach, Leigh Ann Mrotek, Mariah Quinn, Kirsten Abramson, Peter Kleinschmidt, Tom Brazelton, Heidi Twedt, David Kunstman, Graham Wills, John Long, Brian W Patterson, Frank J Liao, Stacy Rasmussen, Elizabeth Burnside, Cherodeep Goswami, Joel E Gordon

## Abstract

**Background:** Ambient artificial intelligence (AI) offers the potential to reduce documentation burden and improve efficiency through clinical note generation. Widespread adoption, however, remains limited due to challenges in electronic health record (EHR) integration, coding compliance, and real-world evaluation. This study introduces a framework and protocols to design, monitor, and deploy ambient AI within routine care.

**Methods:** We launched an implementation phase to build technical workflows, establish governance, and inform a pragmatic randomized trial. A bi-directional governance model linked operations and research through multidisciplinary workgroups that incorporated the Systems Engineering Initiative for Patient Safety (SEIPS) framework. Integration into the EHR used Fast Healthcare Interoperability Resources (FHIR), and a real-time dashboard tracked utilization and documentation accuracy. To monitor drift, a difference-in-differences analysis was applied to three process metrics: time in notes, work outside of work, and utilization. Audits of ICD-10 compliance were performed using an internally-developed large language model (LLM), whose validity was assessed via correlation with certified professional coders.

**Results:** Ambient AI utilization, measured as the proportion of eligible clinical notes completed using the system, had a weighted median of 65.4% (IQR, 50.6% to 84.0%). Iterative improvement cycles targeted task-specific adoption. A brief workflow issue related to a note template change initially reduced ICD-10 documentation accuracy from 79% (95% CI: 72%-86%) to 35% (95% CI: 28%-42%); accuracy returned to baseline after note template redesign and user training. The internally developed LLM coder achieved a strong correlation with professional coders (Pearson’s r = 0.97). The trial enrolled 66 providers across 8 specialties, powered at 90% for the primary outcome on provider well-being.

**Conclusions:** We provide a publicly-available framework and protocols to help safely implement ambient AI in healthcare. Innovations include an embedded pragmatic trial design, human factors engineering, compliance-driven feedback loops, and real-time monitoring to support deployment, ensuring fidelity before initiation of the clinical trial.

**ClinicalTrials.gov ID:** NCT06517082

**DESCRIPTION:** This study presents a comprehensive framework and protocols for safely integrating ambient artificial intelligence (AI) into routine clinical care, focusing on electronic health record (EHR) integration, governance, and real-time monitoring of AI-driven clinical note documentation. The approach includes human factors engineering, compliance audits, and a pragmatic randomized trial design to optimize AI adoption, maintain coding accuracy, and support provider well-being across multiple specialties.

## INTRODUCTION

The rapid commercialization of generative AI has outpaced the development of research methods and regulatory oversight, creating a critical knowledge gap.^1^ This challenge is exacerbated by the fact that health system operations and research organizations often function in silos, creating barriers to evidence-based implementation of care delivery.^2^ Addressing these barriers requires aligning operational and research priorities through structured governance and workflow-aligned evaluation frameworks. Prior studies show that implementing generative AI without tailoring for human factors and organizational contexts often results in underutilization and operational inefficiencies.^1,3^ A Learning Health System (LHS) framework is well-suited to this challenge, providing the structure to bridge the gap between innovation and effective implementation by enabling continuous improvement through iterative, data-driven cycles of evidence generation and use.^4^

We present a pragmatic randomized controlled trial protocol within an LHS framework to evaluate ambient AI software designed to assist with clinical documentation. Clinical documentation in electronic health records (EHR) remains a major contributor to provider burnout, often requiring extensive after-hours work.^5,6^ Although ambient AI aims to reduce this burden, its successful deployment requires complex workflow integration, strict adherence to data privacy, and high documentation accuracy.

Previous studies on ambient AI have demonstrated promising outcomes regarding documentation efficiency and burnout reduction with quality improvement initiatives that incorporate some form of Plan-Do-Study-Act cycles.^7–12^ However, many of these studies were limited by observational designs, a lack of randomization, and short follow-up periods without implementation science frameworks. We address these limitations by providing an adequately-powered, 24-week stepped-wedge randomized trial that employs repeated provider assessments. We integrate implementation science and human factors engineering into our playbook, using Plan-Do-Study-Act cycles,^13^ and frameworks such as the Systems Engineering Initiative for Patient Safety (SEIPS) to ensure organizational alignment and scalability.^14^

This study includes protocols and a framework that we term the Pragmatic Trial Operations Playbook (PTOps), which consists of two interlinked phases for conducting generative AI studies in healthcare. First, the implementation phase focuses on deploying novel data dashboards and monitoring strategies, supported by an integrated governance model bridging operations and research. Second, we describe the design for the clinical trial phase. Our primary hypothesis is that integrating ambient AI into clinical workflows using a bi-directional governance and real-time monitoring framework will yield high adoption within a defined operational timeline and achieve acceptable documentation accuracy and workflow efficiency. The playbook is available in our open-source GitLab repository (https://git.doit.wisc.edu/smph-public/LearningHealthSystem/ambientlistening).

## METHODS

### Pragmatic Trial Operations (PTOps) Playbook

The PTOps playbook consists of five core components: governance, user experience, technical, documentation sustainment (i.e., coding compliance), and analytics. The implementation phase informed the design for the multisite, closed-cohort, stepped-wedge pragmatic randomized clinical trial. The clinical trial protocol followed the Standard Protocol Items: Recommendations for Interventional Trials-Artificial Intelligence (SPIRIT-AI) guidelines.^15^ The protocol and SPIRIT-AI checklist are provided in Supplemental Sections 1 and 2.

### Setting and Environment

Ambient AI (^©^Abridge AI, Inc.) software was integrated into the EHR (^©^Epic Systems, Inc.) under a Software as a Service (SaaS) agreement at the University of Wisconsin Hospitals and Clinics (UW Health). Ambient AI use was part of a quality improvement initiative, with patients providing informed consent during routine care, while providers participating in the clinical trial consented to the research component. Since ambient AI licenses were purchased for clinical use regardless of the trial, the study qualified for expedited review under 45 CFR 46.110 by the UW Institutional Review Board (UW IRB #2024-1028), posing no more than minimal risk. A stepped-wedge design was selected due to the impracticality of rolling out the intervention to all providers simultaneously. The design also allowed for random concurrent exposure to isolate the ambient AI system’s effects from secular trends while maintaining guaranteed access for all providers as part of the clinical operations strategy.^16,17^

### PTOps Playbook Part A: Governance

The initial governance model effectively guided the implementation phase and informed the design of the clinical trial. As the transition into the clinical trial phase was made, the operations team updated and refined the governance structure to support ongoing decision-making and scalability. The updates are shown in Figure 1, which is divided into two parts: Figure 1A illustrates the initial implementation phase with LHS integration and the development of playbook components for evaluation and monitoring. Figure 1B shows the extended governance model applied during the clinical trial phase, including an Escalation Matrix (Supplemental Section 3) for managing decisions on key performance indicators and software license allocation.

**Figure 1.**
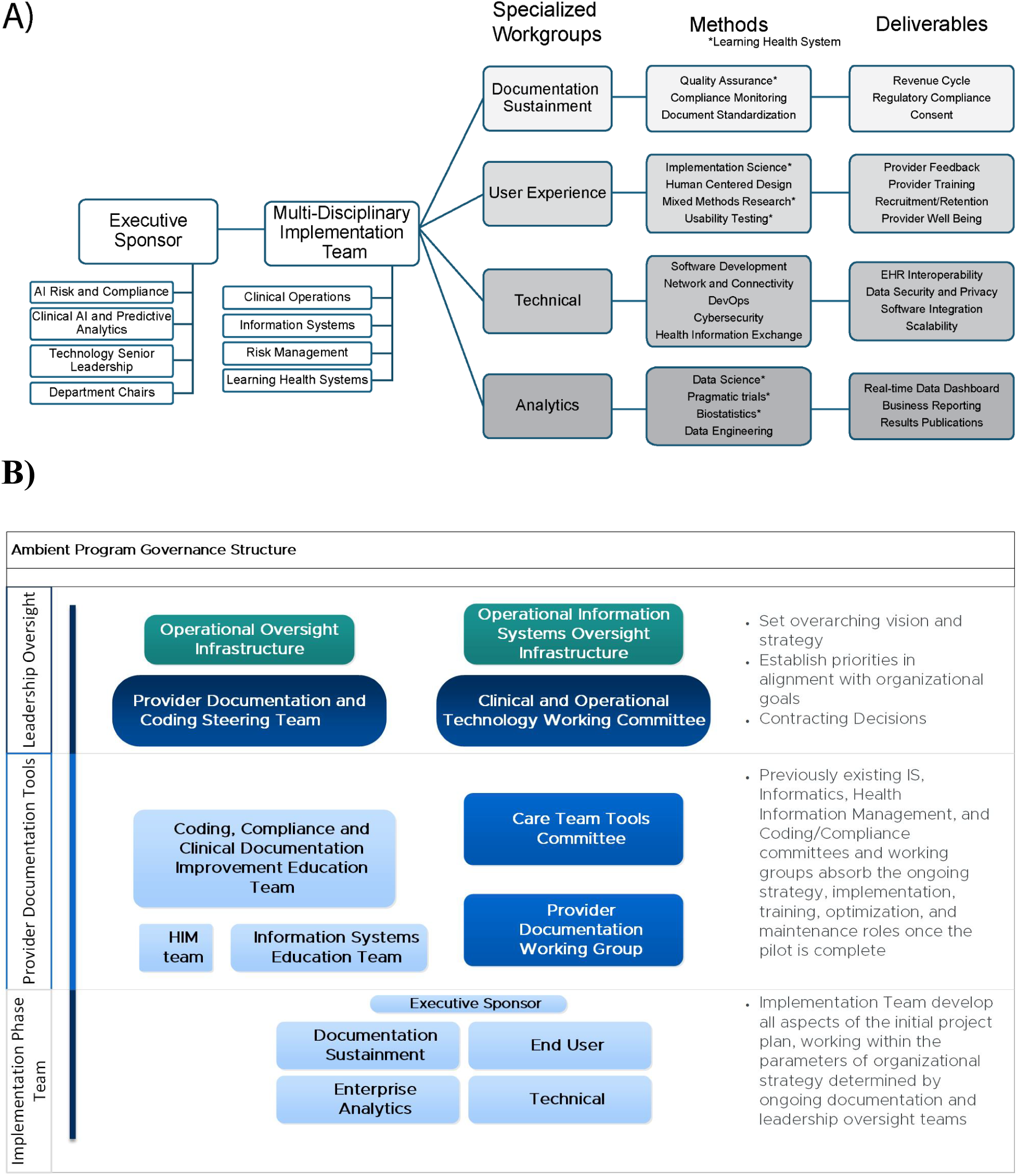
Governance across the initial Implementation Phase with Learning Health System integration (1a) and subsequent design for the Clinical Trial Phase for Operations (1b) Figure 1a: Displays the integration of LHS services (denoted by an asterisk) and outlines the deliverables that informed the playbook components establishing evaluation and monitoring strategies. Figure 1b: Presents the refined operational governance model derived from the implementation phase, to be applied during the clinical trial and subsequent clinical operations. EHR = electronic health record; HIM = health information management.

### PTOps Playbook Part B: User Experience

User experience was informed by two frameworks: the Exploration, Preparation, Implementation, and Sustainment (EPIS)^18,19^ and modified Systems Engineering Initiative for Patient Safety (SEIPS).^14^ The EPIS framework highlighted the interplay of contextual factors, including outer context (organizational readiness), inner context (provider characteristics), innovation (ambient AI), and bridging factors (communication and coordination). The SEIPS model, with its People, Environment, Tools, and Tasks (PETT) Scan,^20^ offered a practical approach for mapping work processes and identifying potential barriers and facilitators (Supplemental Section 4). Training materials and workflows were iteratively refined based on provider feedback in REDCap surveys^21^ with Plan-Do-Study-Act cycles.

### PTOps Playbook Part C: Technical Integration and Onboarding

Integration leveraged Epic’s private Application Programming Interfaces (APIs) and Fast Healthcare Interoperability Resources (FHIR) R4 APIs to transfer AI-generated notes and session metadata securely into the EHR. Providers obtained verbal consent from patients and used the Epic Haiku mobile application to initiate recordings (Figure 2). Key prerequisites included importing the Abridge client record (which contains FHIR credentials such as client IDs) and collaborating closely with Epic Technical Support. The technical workgroup coordinated system access and deployment, aligning these efforts with the clinical trial’s randomization schedule, while daily reviews ensured consent compliance and system performance. The integration process involved configuring the Interconnect system to allow Abridge API calls, setting up outgoing message queues and listeners, and establishing OAuth2 authentication.

**Figure 2.**
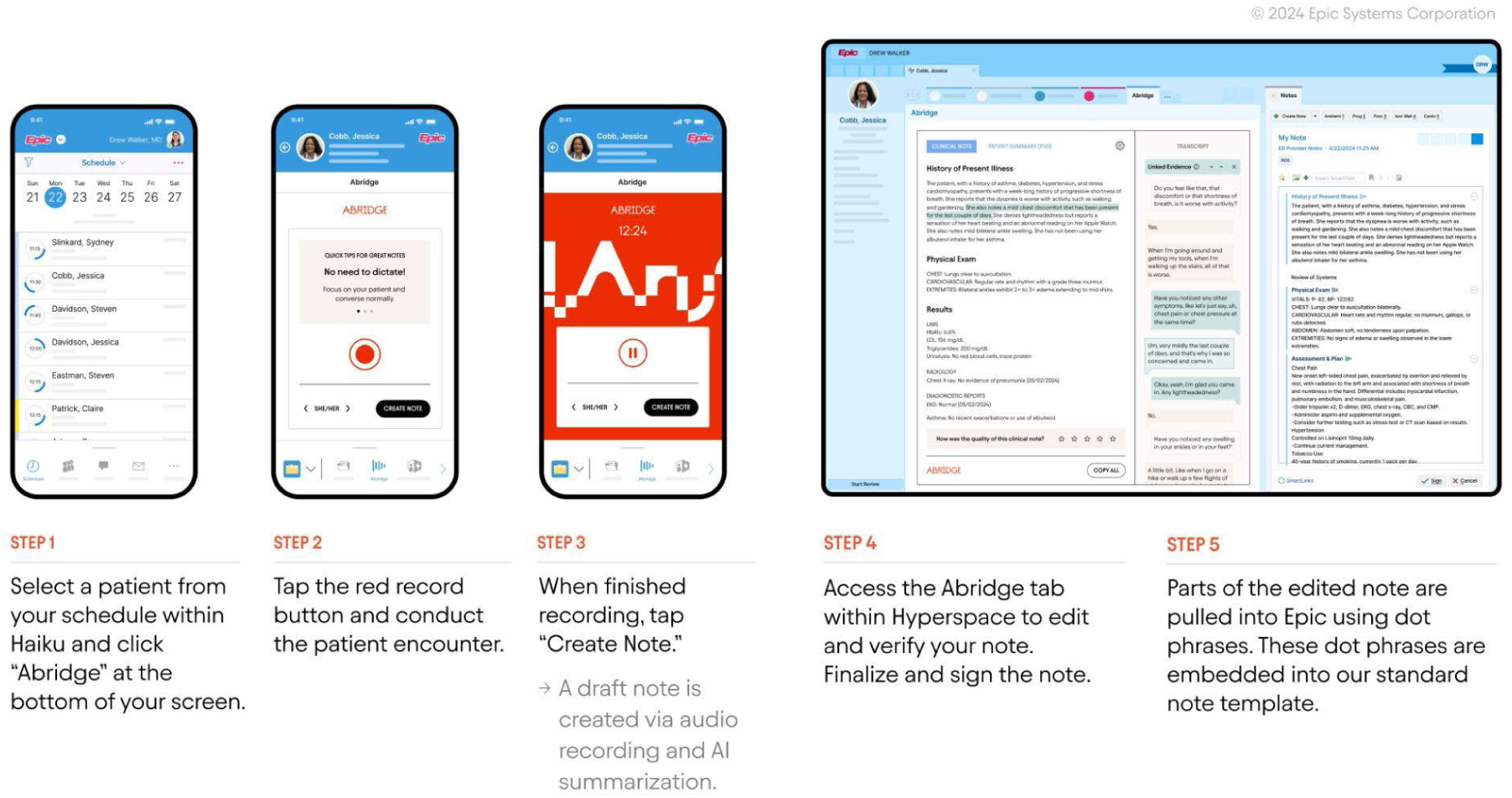
**Abridge ambient software integration with Epic electronic health record system.**

For the clinical trial design phase, the inclusion criteria were providers treating at least 20 patients per week, completing the required training, and using Epic Haiku. Providers with planned leave, unsupported mobile devices, or unwillingness to discontinue existing medical scribes were excluded. Operational staff coordinated with the research team to contact eligible providers, schedule training sessions, and oversee technical onboarding.

### PTOps Playbook Part D: Process Measures and Data Dashboard Monitoring

A real-time data dashboard (^©^1993-2024 QlikTech International AB) integrated data from the EHR and Abridge software to monitor process measures (Supplement 5).^22^ Dashboard metrics included provider feedback, utilization rates, patient consent compliance, and documentation efficiency. This last metric was quantified by work outside of work (WoW) and time spent on notes (both normalized to the 8-hour workday) as well as closures before next encounter by end of day, and patient follow ups.^22^ Since licenses were issued at the provider level, the weighted medians of average daily provider-level metrics were measured to capture central tendencies and variability.^25^ Weights were created by a provider’s relative number of in-clinic days compared to the total number of days.

An LLM for ICD-10 code extraction from AI-generated notes was developed and evaluated internally at UW Health. The evaluation focused on ensuring accurate identification of the 10 most frequent diagnosis codes. We deployed OpenAI’s GPT-4o LLM within a secure, HIPAA-compliant Azure environment to generate ICD-10 codes from clinical documentation. Prompt design incorporated four prompt engineering strategies: minimizing perplexity, in-context examples, chain-of-thought reasoning, and self-consistency.^24–26^ Evaluation metrics included precision, recall, and macro-averaged F1-score with 95% confidence intervals (CI) based on 100 bootstrapped samples. In the operational setting, LLM-derived documentation compliance scores were compared against those assigned by certified professional coders. Pearson correlation was used to assess agreement. Thresholds for actionability were informed by professional coder review and implemented using a zone-based Escalation Matrix (Supplemental Section 3). Detailed prompt construction and error definitions are described in Supplemental Section 6.

A difference-in-differences (DiD) analysis was implemented to detect workflow issues in near real-time, arising from the interaction between the AI software, EHR, and human workflows. Drift was assessed across three process metrics identified by Information System stakeholders as most sensitive to workflow issues: (1) time in notes; (2) work outside of work; and (3) utilization. Observations were collected in one-month windows, updated every two weeks. A unique feature of this approach was that the proportion of providers exposed to ambient AI increased over time, which changed the distribution of observations between arms. When both ambient AI and usual care (trial-enrolled providers who had not yet been given access to ambient AI) data were available in a window, the difference in between-arm changes were compared for each metric from the first two weeks to the last two weeks. When only one condition was present, within-group changes over time were analyzed. This DiD system was piloted during the implementation phase and tested during a major note template change as a real-world use case. Analyses employed linear mixed-effects models with random intercepts for individual providers and fixed effects for time, accounting for autocorrelation across two-week intervals. Actionable drifts were identified as those differences with Bonferroni corrected p-values ≤0.05, adjusted for the total number of tests performed. Identification of such drifts triggered root-cause analyses and discussions with operational leaders.

The clinical trial phase was designed as a 24-week, individually randomized, stepped-wedge trial. In collaboration with our Chief Wellness Officer, the Professional Fulfillment Index (PFI)^27^ — a validated provider survey routinely administered at UW Health to measure well-being — was defined as the primary outcome. Historical PFI survey data from 1,091 providers at UW Health in 2023 informed the clinical trial’s power and sample size calculations. Providers were randomized 1:1:1 into three waves using stratified permuted-block randomization, ensuring balance across clinical specialties.

## RESULTS

The implementation phase started on June 24, 2024, until the initiation of the clinical trial on October 10, 2024. Twenty providers with 8,527 clinic encounters were evaluated during the implementation phase. The providers were distributed across 5 specialties, 12 clinic locations, and 50% were female. Utilization rates were analyzed to determine a minimum acceptable threshold for ambient AI use, with the lower limit set at 48%, defined as three standard deviations below the mean. This threshold guided license allocation and fidelity tracking by the operations team. During the implementation phase, the weighted median of provider-averaged daily ambient AI utilization was 65.4% (IQR: 50.6% - 84.0%). The average time in notes decreased by 0.35 hours (95% CI: 0.05 – 0.64) in the final two weeks compared to the first two weeks.

Ten providers (50%) participated in interviews to share feedback about their experience with ambient AI. Positive comments included statements such as, “I feel like I was walking before, and now this is like a bullet train, and a scribe would be something like a stagecoach.” Providers also highlighted challenges, such as, “I have had some patients decline, citing privacy concerns even after discussing how it works and that it’s compliant.” Key themes emerged from the interviews: (1) organizational characteristics, including clinic-level physical environments and team dynamics, influenced ambient AI utilization; (2) provider and patient readiness, as well as documentation preferences, contributed to variability in adoption; and (3) the suitability of ambient AI platforms to offload providers’ documentation burden depended on service settings and clinical tasks. Interviewers addressed these barriers and identified recommendations (Table 1). Plan-Do-Study-Act cycles were employed to address issues such as inconsistent use of note templates and gaps in obtaining patient consent. These iterative modifications included updating training materials, enhancing technical support, and refining workflows.

**Table 1.** Key Themes mapped to SEIPS PETT Scan with Examples of Barriers and Facilitators to Inform Implementation (B= Barriers; F = Facilitators)

| Theme | People | Environments | Tools | Tasks |
| --- | --- | --- | --- | --- |
| 1. Organizational characteristics | -Time pressure (B)<br>-Documentation pressure/stress (BF) | -Diverse physical settings (B)<br>-Centralized implementation team (F)<br>-Standardized workflows (F) | -Internet/server connectivity issues (B) | -Technical assistance (BF) |
| 2. Provider and patient readiness (individual characteristics) | -Understanding, willingness, and learning curve (B)<br>-Provider documentation preferences (B)<br>-Excited about new technology (F) | -Preferences for specific clinics (B)<br>-App burden (B) | -Quirk tolerance for tool (BF) | -More connection and eye-contact with patients (F) |
| 3. Type of service setting and clinical task needs | -Acceptability/usability of tool and generated note (BF) | -Templates for state specific requirements (B) | -Templates for specialties & types of visits (B)<br>-Technology quirks/glitches (B)<br>-Integration with her (F) | -Not able to do certain tasks (pending orders, pre-visit summaries) (B)<br>-Compliance issues (B) |
| Suggested Implementation Strategies and Adaptations (EPIS phase)<br>• Audit and provide feedback (IS)<br>• Booster training sessions (PIS)<br>• Centralized and local technical assistance (PIS)<br>• Clear guidance (PIS)<br>• Communicate data to external stakeholders, providers, and patients to demonstrate ongoing benefit (IS)<br>• Information for patients (PIS)<br>• Positive messaging and testimonials about value (EPIS)<br>• Stage implementation scale-up (PI)<br>• Tools for quality monitoring (PIS)<br>• Training materials and FAQ sheets for providers (PIS)<br>• Training and reminders for compliance (IS) |  |  |  |  |

A manual review of 797 visit encounters across 17 providers revealed an issue during initial workflow integration. In the pre-ambient note template, all visit diagnoses entered during the encounter by the provider, including diagnoses attached to orders or medications, were automatically inserted into the provider’s standard clinic note template as a reminder to include supporting documentation, promoting consistency between coded diagnoses and clinical notes. Post-intervention, the display was not included in the note template during the initial workflow integration efforts in an attempt to simplify the note template and leverage the content of the AI generated assessment and plan, resulting in a drop in diagnostic coding accuracy from an average of 79% (95% CI: 72% - 86%) to 35% (95% CI: 28% - 42%); We identified three types of discrepancies: (1) diagnoses present in the note but omitted from the visit diagnosis list, which are needed for code justification; (2) diagnoses included in the visit list that were not documented in the note, increasing the risk for claims denials; and (3) mismatches in specificity, where less detailed codes replaced more specific ones, potentially degrading complexity measures (e.g., Hierarchical Condition Categories). To mitigate these issues — reflecting an EHR workflow challenge external to the core ambient AI process but requiring integration for resolution — we introduced a refreshable block of text (i.e., SmartLink in Epic) into a revised standard note template that automatically populated the list of visit diagnoses along with supporting compliance information like consent by the patient for ambient use (Supplemental Section 7). Targeted training and updated onboarding procedures further improved documentation, with accuracy returning to pre-intervention levels of an average 80% (95% CI: 76% - 84%).

To support near real-time data safety monitoring, a DiD analysis was conducted across 13 rolling windows. In examining a 4-week window when the note template workflow issue surfaced, there was an average decrease in ambient AI utilization of 6.84 percentage points (95% CI: -18.50 – 4.82), an average increase in work outside of work by 0.33 hours (95% CI: -1.18 – 1.89), and an average increase in time spent in notes by 0.43 hours (95% CI: -0.05 – 0.91) (Figure 3).

**Figure 3.**
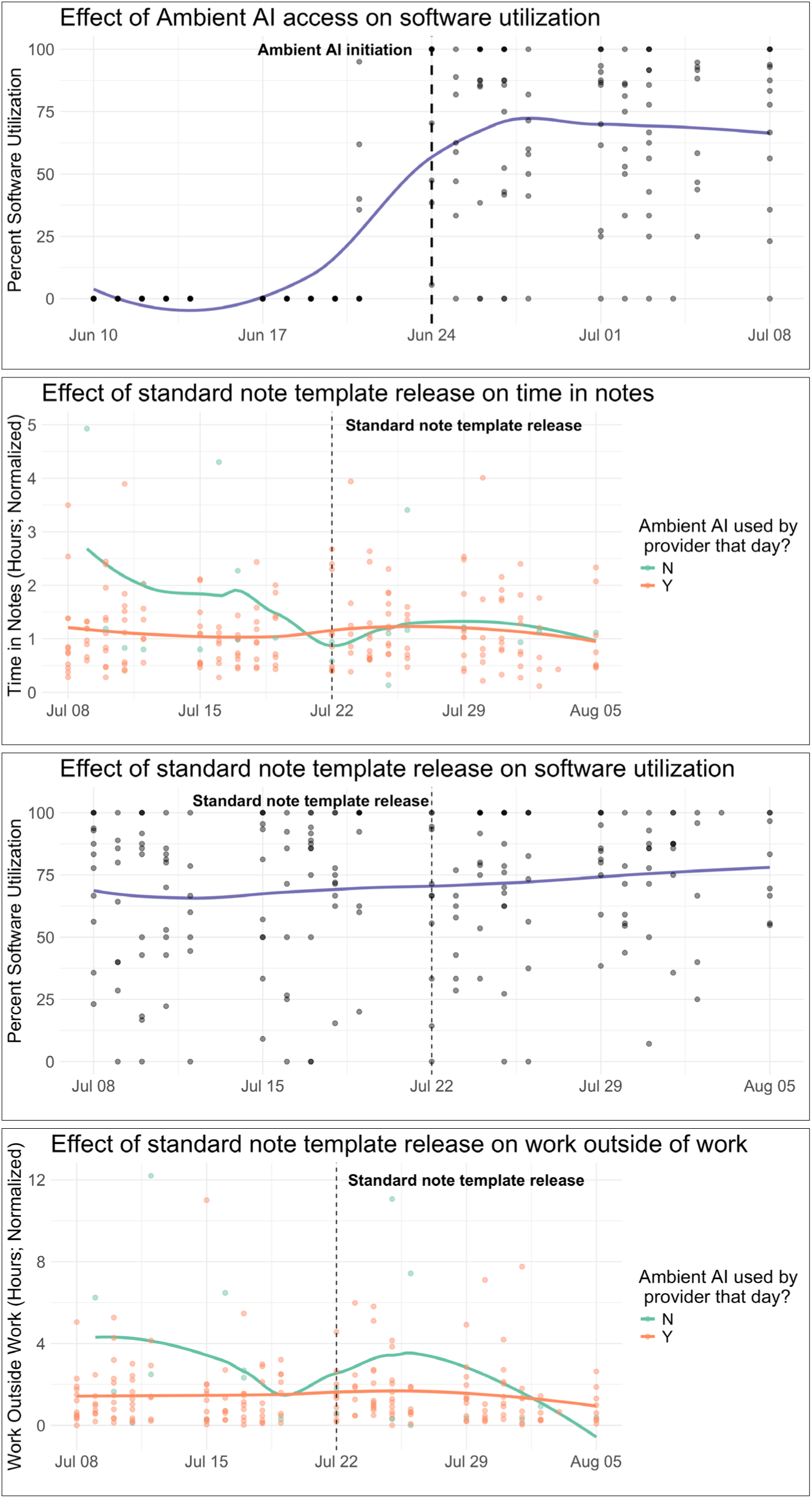
Differences-in-differences across 4-week window for ambient AI initiation and new standard note implementation Differences-in-differences across 4-week window for ambient AI initiation and new standard note implementation. Panel 1 illustrates ambient AI utilization rates in the two weeks before and after users were given full access to the software. Panels 2-4 pertain to the two weeks before and after a new standard note template was implemented, depicting utilization rates, time in notes, and work outside of work for providers in this period. Each dot represents an observation for a provider on a given working day. For panels 1 and 2, the blue line represents a loess-smoothed trend line in ambient AI utilization rates. For panels 3 and 4, the green line represents the loess-smoothed trend line for time in notes for days where a provider did not use ambient AI; the orange line represents the loess-smoothed trend line for time in notes for days where a provider did use ambient AI. In each panel, the vertical line indicates the date of a major event: for panel 1, this is the date on which most of the providers were given access to ambient AI software; for panels 2-4, this is the date on which the new standard note implementation went into effect.

Professional coders logged a total of 598.33 hours manually reviewing diagnosis documentation accuracy. Recognizing the unsustainability of manual review for the larger cohort of providers in the clinical trial phase, an automated LLM coding review system was developed. A total of 30,468 encounters over three months were analyzed, focusing on the top 10 most frequent ICD-10 diagnoses. The median input word count to the LLM coder was 734 words (IQR: 560– 998). When compared with professional coders, the LLM coder achieved a macro-averaged precision of 0.88 (95% CI: 0.87–0.88), a recall of 0.66 (95% CI: 0.65–0.67), and an F1-score of 0.73 (95% CI: 0.72–0.73). The median diagnosis compliance scores were 9.0 (IQR: 5.0–9.0) for professional coders and 8.4 (IQR: 2.0–9.3) for the LLM coder. The correlation of the compliance scores between the LLM coder and professional coders was strong, with a Pearson’s r of 0.97 (95% CI: 0.97–0.98).

By October 2024, the recruitment of the planned 66 providers across eight specialties was completed. The clinical trial began on schedule, and the randomization achieved balanced allocation across the waves. In Wave 1, 45.5% of providers were from Family Medicine, 27.3% from Internal Medicine, 9.1% from Pediatrics/Adolescent, and 18.2% from Miscellaneous/Other; Wave 2 comprised 42.9%, 28.6%, 9.5%, and 18.9% respectively; and Wave 3 included 47.8%, 26.1%, 8.7%, and 17.4% in the same order. Post-implementation, the data dashboard monitoring system transitioned to University of Wisconsin Hospitals and Clinics (UW Health) as the owner, ensuring operational stewardship.

### Protocols and Study Documentation

To promote transparency and reproducibility, all protocols, study documentation, and analysis code were released through a public GitLab repository. The repository is organized into four sections: (I) *Governance and Regulatory*, including the IRB application and approval materials, the clinical trial protocol, randomization code, and data dictionary; (II) *User Experience*, encompassing pre-implementation, implementation, and trial-phase interview guides, survey instruments with codebooks and results, and implementation frameworks (EPIS and SEIPS); (III) *Technical Integration and Onboarding*, including integration documentation, a user guide, and clinician-facing materials such as flyers and tips for use with trainees; and (IV) *Analytics and Data Dashboard Monitoring*, which provides analytic code and documentation for DiD analysis, LLM evaluation, and real-time dashboard metrics (https://git.doit.wisc.edu/smph-public/LearningHealthSystem/ambientlistening).

## DISCUSSION

This study demonstrates how aligning research and operational priorities can accelerate the implementation and evaluation of rapidly evolving generative AI technology. A key to success was the governance structure, which supported iterative improvement cycles.^28^ While prior studies have shown promise for ambient AI, most have relied on observational designs and lacked generalizable implementation frameworks to guide other health systems.^8–12^ In contrast, we employed a randomized controlled trial embedded within a pragmatic, real-world setting, allowing for causal inference on outcomes such as documentation efficiency, provider burnout, and patient experience. In addition, our longitudinal monitoring strategy incorporates real-time analytics to track performance, user feedback, and performance drift. These efforts aligned with three key characteristics of socio-technical infrastructure:^29^ (1) engaging multi-stakeholder learning communities; (2) rigorously exploring uncertainty during the implementation phase; and (3) fostering sustainability through iterative improvement cycles.

Informatics innovations included automating ICD code review with LLMs and integrating real-time data feeds from the EHR into an operational dashboard. These tools reduced reliance on manual data collection and supported a scalable evaluation model. Another key innovation was adapting trial designs to the dynamic nature of generative AI systems. Unlike static interventions in traditional trials, generative AI platforms continue to have updates that may lead to significant drifts in performance. The lack of adverse event categories for what constitutes major or minor AI software changes necessitated a monitoring strategy to detect unexpected drifts during deployment. While our DiD analysis initially showed a modest increase of approximately 25 minutes for time in notes following a new note template workflow, reflecting transitional workflow adjustments, the subsequent paired analysis indicates an overall reduction in documentation time by approximately 20 minutes for time in notes as providers adapted to the improved template and refined their workflows.

Operationalizing ambient AI use required more than a simple “plug-and-play” implementation. Our experience shows that a multidisciplinary governance structure — encompassing technical integration, documentation compliance, and clear decision rules based on analytics — helped achieve high adoption and maintain coding integrity. Functioning as an analog to a Data Safety and Monitoring Board, our dashboard addressed both the immediate needs of the trial and the long-term goals of monitoring and scalability. Our human-centered approach and in-depth operational monitoring yielded higher utilization rates than those observed in prior studies.^8–12^ Weekly reviews allowed targeted outreach — via low-utilizer notifications and personalized mentoring — to optimize license usage and refine integration strategies. This iterative feedback loop was important in aligning the technology with provider practices, thereby enabling effective implementation. To facilitate broader adoption, we have made our protocols and comprehensive playbook publicly available at https://git.doit.wisc.edu/smph-public/LearningHealthSystem/ambientlistening. The repository includes detailed regulatory documentation, onboarding materials, monitoring and analytics code, REDCap surveys, trial protocol with randomization code, and operational resources that outline the entire process from implementation to ongoing trial management.

Several limitations are worth noting. Participation in the trial may skew toward providers with higher technological literacy or interest in adopting new tools, introducing a selection bias. While this may limit generalizability to all providers, it reflects the intended user base for software license distribution in real-world settings, where uptake is often driven by interest and readiness. There is also potential for a Hawthorne effect, as asking providers to complete the PFI may heighten their awareness of specific aspects of their work, thereby influencing how they perceive or report their experiences. Also, knowing that clinical encounters are being recorded and transcribed may alter provider behavior. While the former is likely to have minimal impact due to the longitudinal nature of the trial, the latter may be more pronounced early in the intervention period. However, any sustained behavioral change related to awareness of being recorded may be considered part of the overall intervention effect. Unlike prior studies with cross-sectional designs, our trial allows for the evaluation of time-varying treatment effects.

As generative AI technologies continue to evolve rapidly, it is crucial to deploy, monitor, and adapt implementation in real time. We provide an open-source playbook for bridging the gap between innovation and real-world application, helping to support the safe, effective, and sustainable adoption of AI in clinical care.

## Data Availability

All data produced in the present study are available upon reasonable request to the authors.

## ACKNOWLEDGEMENTS

Tom Wise (Business Relation Management), Michele L Nickels, Christine Cunningham (Health Information Management), Troy Lepein (Risk and Compliance/Business Integrity), Karen Nachman, Luke Rislove and Sarah Fardy (Enterprise Analytics), Rachelle Buol and Tori L McKinley (Clinical Documentation Integrity), Nicole Riechers (IS Project Management Office), Lisa Wilson and Monica Esquibel (Institutional Review Board), and Betsy Nugent and Nasia Safdar (Clinical Trials Institute).

